# Increased levels of anxiety among medical and non-medical university students during the COVID-19 pandemic in the United Arab Emirates

**DOI:** 10.1101/2020.05.10.20096933

**Authors:** Basema Saddik, Amal Hussein, Fatemeh Saheb Sharif-Askari, Waad Kheder, Mohamad-Hani Temsah, Rim Adnan Koutaich, Enad Sami Haddad, Nora Marwan Al-Roub, Fatema Adel Marhoon, Qutayba Hamid, Rabih Halwani

## Abstract

**Introduction:** The COVID-19 pandemic is likely to increase anxiety levels within the community and in particular medical students who are already considered psychologically vulnerable groups. Since the COVID-19 outbreak, no study has yet estimated the effect of this pandemic on university students in the UAE or its impact on the psychological well-being of medical students.

**Methods:** In this study, we surveyed 1485 medical (comprising medical and dental) and non-medical university students across 4 emirates within the UAE. We used an online platform to assess knowledge, sources of information, changes in hygienic behavior, perceptions of fear and worry and anxiety levels using the generalized anxiety disorder 7 (GAD-7) scale. The GAD-7 score was measured at three time points; during hospital visits for medical/dental students, before the introduction of online learning and after online learning for all students.

**Results:** The majority of students demonstrated high levels of knowledge and utilized reliable sources of information. Non-medical students exercised higher compliance with social restrictions, while medical students practiced better hand hygiene. Almost half of students reported anxiety levels ranging from mild to severe with females reporting higher anxiety scores during hospital visits (OR=2.02, 95% CI, 1.41 to 2.91) and medical students reporting lower anxiety levels in comparison to dental students (OR=0.61, 95% CI, 0.45 to 0.84). Medical students reported higher levels of anxiety during their clinical rotations which decreased with the introduction of online learning, yet, non-medical student’s anxiety levels increased with online learning.

**Conclusions:** This is the first study to provide important information on the initial response and anxiety levels in university students across the UAE. The findings from our study can be used to support the development of effective screening strategies and interventions to build psychological resilience among university students during the COVID-19 pandemic or any other public health emergencies in the future.

## Introduction

Human coronaviruses (HCoVs) have long been considered inconsequential pathogens, causing the common cold in otherwise healthy people. However, in the 21st century, two highly pathogenic HCoVs—severe acute respiratory syndrome coronavirus (SARS-CoV) and Middle East respiratory syndrome coronavirus (MERS-CoV)—emerged from animal reservoirs to cause global epidemics with alarming morbidity and mortality in 2003 and 2012 respectively. In December 2019, yet another pathogenic HCoV, SARS-CoV-2, was recognized in Wuhan, China, causing Coronavirus disease (COVID-19) [1] and raising intense attention not only within China but internationally [1].

Since its first detection, COVID-19 has become a major health problem and the global outbreak was declared a pandemic by the World Health Organization (WHO) on March 11^th^, 2020 [2]. To date, more than 3.4 million laboratory-confirmed cases have been reported worldwide with ~245,000 deaths in 184 countries, bringing the crude case fatality rate (CFR) of COVID-19 to ~7.2%[3]. The first confirmed case of COVID-19 in the United Arab Emirates (UAE) was reported on 29 January 2020. The UAE had 13,599 confirmed cases (10,816 active; 2,664 recovered; 119 deaths; CFR ~1.1%, as of 3^rd^ May 2020) [4].

The emergence of such a novel and highly contagious pandemic and its uncertainty is likely to raise community anxiety and have a psychological impact on communities overall. Social distancing, quarantine and disruption of studies, are especially pertinent on impacting university students, and in particular medical students who are already considered psychologically vulnerable groups. The highly competitive training of these students, their academic pressure, exposure to patients in clinical settings, financial constraints and lack of sleep are factors which may already contribute to psychological problems associated with stress and anxiety [5-7]. Additionally, during disease outbreaks, medical students, like healthcare workers, are perceived to be at higher risk of infection because of their clinical training, and hence increased risk of exposure to the virus [8, 9]. This is in addition to their fear of transmitting the virus back to their family and loved ones [10].

In previous infectious disease outbreaks, such as MERS-CoV and SARS, almost one quarter of medical students reported mild to moderate anxiety in Saudi Arabia [11] and anxiety levels were found to be higher in medical students than non-medical students in Hong Kong [12].

Since the COVID-19 outbreak, no study has yet estimated the effect of this pandemic on university students in the UAE and its impact on the psychological well-being of medical students. In this study, we aim to assess medical student’s psychological distress and concerns during the recent COVID-19 pandemic, their degree of perceived information about the disease, and their overall attitude, practices and behaviors during the outbreak. We also evaluate students’ perceptions on the precautionary measures in place and the effectiveness of educational strategies in reducing anxiety levels in the United Arab Emirates. This makes our study the first in the UAE to discuss this aspect in the current pandemic.

## Methods and Materials

### Design and participants

A cross-sectional study design was used to conduct this research during the COVID-19 outbreak in March 2020. Data were collected between March 11 and March 21. University students from medical, dental and non-medical colleges across the UAE were invited to participate in the research through an online web survey hosted on the platform survey monkey (https://www.surveymonkey.com/r/VSYTNQ9)[13] (S1). All students enrolled in the medical and dental colleges at the University of Sharjah received the survey link through their university emails using convenience sampling. Students from other universities received the survey link through WhatsApp and other social media platforms using snowball sampling where acquaintances sent the link to each other. Through the survey link, the first page explained the research objectives and assured participants anonymity and confidentiality. Students’ acceptance indicated their consent to participate in the study. The study was approved by the University of Sharjah Research Ethics Committee (REC-20-03-03-02) prior to participant recruitments.

### Sample size

A study conducted in Saudi Arabia during the MERS-COV outbreak found that 77% of medical students reported minimal anxiety levels [11]. Using this proportion and a confidence level of 95% and margin of error of 5%, we calculated the minimum required sample size to be 278 for each of the medical and non-medical groups. To account for non-response the sample size was increased by 20%, making the minimum sample size required for this study 333 students for each of the medical and non-medical groups.

### Data collection

A questionnaire comprising of 18 items was used in this study. A modified version of a questionnaire measuring medical student anxiety previously used in Saudi Arabia during the MERS-CoV outbreak was used, after the author’s permission[10, 11]. The questions were divided into seven domains comprising of 1) demographic questions, 2) change in hygienic behavior, with responses measured on a 4 point Likert scale ranging from very much has changed to no change at all, 3) level of knowledge on statements related to COVID-19 such as transmission, treatment, prognosis and prevention measured by true/false/don’t know responses, 4) perceptions of worry and fear associated with COVID-19 measured on a 5-point Likert scale, ranging from very worried to not worried at all, and their opinion about the public fear associated with COVID-19 being justifiable or dysfunctional, measured on a 5-point Likert scale, ranging from strongly agree to strongly disagree, 5) perception of receiving enough information on COVID-19 on a 5-point Likert scale, ranging from strongly agree to strongly disagree, 6) the sources of information students resorted to for information on COVID-19 and 7) Measurement of anxiety levels.

In order to measure anxiety levels, we used the generalized anxiety disorder scale (GAD-7) [14] which is a 7-item questionnaire asking participants how often they were bothered by each symptom such as feeling nervous, trouble relaxing, irritable and afraid that something awful might happen during the last 2 weeks. Response options were “not at all,” “several days,” “more than half the days,” and “nearly every day,” scored as 0, 1, 2, and 3, respectively. A score of 10 or greater represents a reasonable cut point for identifying cases of anxiety with a sensitivity of 89% and specificity 82%, internal consistency (Cronbach α=.92) and Test-retest reliability (intraclass correlation=0.83)[14]. The GAD-7 has also been identified as a screener for panic disorder, social phobia and PTSD (with a cutoff score of 8 sensitivity 77% and specificity 82%) [15]. In our study, the GAD-7 score was totaled for each student and classified into cut-off points of (0-4 minimal, 5-9 mild, 10-14 moderate and 15-21 severe) levels of anxiety. Levels of anxiety were measured using the GAD-7 scale at three different time points. In addition to calculating the mean GAD-7 score, scores were classified into 3 categories (minimal, mild and moderate/severe anxiety).

During the launch of data collection for this study, the UAE Ministry of Education declared university campuses to move to online learning to prevent the spread of COVID-19 and classes moved online on March 8. In order to determine levels of anxiety of students during the transition to online learning, medical and dental students were asked to respond to the GAD-7 questions reflecting back to the time they were attending hospital visits within the previous two weeks. This was to assess whether higher levels of anxiety were associated with hospital visits. All students were asked to respond to the GAD-7 questions reflecting on “before the introduction of online learning”, and “after the introduction of online learning”.

The final version of the questionnaire was piloted to ensure clarity and consistency between survey items. To ensure face and content validity of the survey instrument, the survey was sent to a group of 9 experts which consisted of students, tutors, faculty and a psychiatrist who reviewed the survey for content accuracy, clarity and comprehensiveness and whether the survey met its objectives. As a result, the phrasing and response items of some questions were modified, and format was edited for clarity and comprehensibility

### Statistical analysis

Descriptive statistics, including means, medians, frequencies and percentages were used to summarize data and to illustrate the demographic and other selected characteristics of students. Normality of data was tested visually using the Q-Q plots and statistically using the Kolmogorov-Smirnov test. Bivariate analysis using Chi-square (χ^2^) and Mann-Whitney U tests explored the associations between student demographic characteristics and anxiety levels. Spearman’s correlation coefficient, r, was used to evaluate the association between knowledge score and GAD-7 score. Statistically significant factors in the bivariate analysis were included in the multivariate ordinal logistic regression analyses to determine if they predicted student anxiety levels. The estimates of the strengths of associations were demonstrated by the odds ratio (OR) with a 95% confidence interval (CI). A two-tailed *p <0.05* was considered statistically significant. Data were analyzed using IBM SPSS Version 25.0 [16]

## Results

In total, 1484 students responded to the questionnaire and data were analyzed for 1385 completed surveys (completion rate 93.3%) from 4 different emirates across the UAE. The mean age of students was 20 years and most participants were females (72%). Almost three quarter of the students were studying medicine or dental medicine and from those students a third (35%) were in their clinical years of study and were completing clinical ward rotations. Of these students, (12%) reported being in contact with an infected or suspected case of COVID-19. Demographic characteristics are shown in Table 1.

**Table 1:** Demographic characteristics of students (N=1385)

|  |  | Frequency (n) | Percentage (%) |
| --- | --- | --- | --- |
| **Age (years) [mean ± SD]** | 20.5 ± 2.3 |  |  |
| **Gender** |  |  |  |
| Female |  | 994 | 71.8 |
| Male |  | 391 | 28.2 |
| **Emirate** |  |  |  |
| Sharjah |  | 814 | 58.8 |
| Dubai |  | 107 | 7.7 |
| Ajman |  | 446 | 32.2 |
| Al-Ain/RAK |  | 18 | 1.3 |
| **Field of study** |  |  |  |
| Medical |  | 719 | 51.9 |
| Dental |  | 323 | 23.3 |
| Non-medical |  | 343 | 24.8 |
| **Phase of study for medical/dental students** |  |  |  |
| Pre-Clinical |  | 685 | 65.5 |
| Clinical |  | 361 | 34.5 |
| **Current ward rotation for clinical students\*** |  |  |  |
| Not in a rotation |  | 33 | 9.8 |
| High risk rotation |  | 25 | 7.4 |
| Low risk rotation |  | 280 | 82.8 |
| **Contact with suspected COVID-19 patient** |  |  |  |
| Yes |  | 152 | 12.1 |
| No |  | 1109 | 87.9 |
| **GAD-7 Score Median [IQR]** |  |  |  |
| *During hospital visits (medical/dental students)* | 4 [8] |  |  |
| Minimal |  | 368 | 53.1 |
| Mild |  | 166 | 24.0 |
| Moderate |  | 82 | 11.8 |
| Severe |  | 77 | 11.1 |
| *Before online learning (all students)* | 4 [9] |  |  |
| Minimal |  | 570 | 52.3 |
| Mild |  | 274 | 25.1 |
| Moderate |  | 116 | 10.6 |
| Severe |  | 130 | 11.9 |
| *After online learning (all students)* | 3 [8] |  |  |
| Minimal |  | 620 | 56.9 |
| Mild |  | 254 | 23.3 |
| Moderate |  | 105 | 9.6 |
| Severe |  | 111 | 10.2 |
| **Knowledge Score Median [IQR]** | 5 [1] |  |  |

\*High risk rotation includes ICU, ER, OR, Isolation wards

### Hygienic Practices

The majority of students reported increased hand hygiene (85%), increased use of hand sanitizer (85.5%), avoiding people with flu like symptoms (80%) and decreased visits to crowded places (77%). A lower proportion of students reported wearing gloves and masks (58%), decreased social visits (62%) or decreased hand shaking (58%). We combined medical and dental students together as the medical group, and all other specialties made up the non-medical group. When medical and non-medical students were compared, more non-medical students avoided being in contact with people with flu-like symptoms, had decreased social visits, decreased visits to crowded places and use of public facilities as displayed in Figure 1.

**Figure 1:**
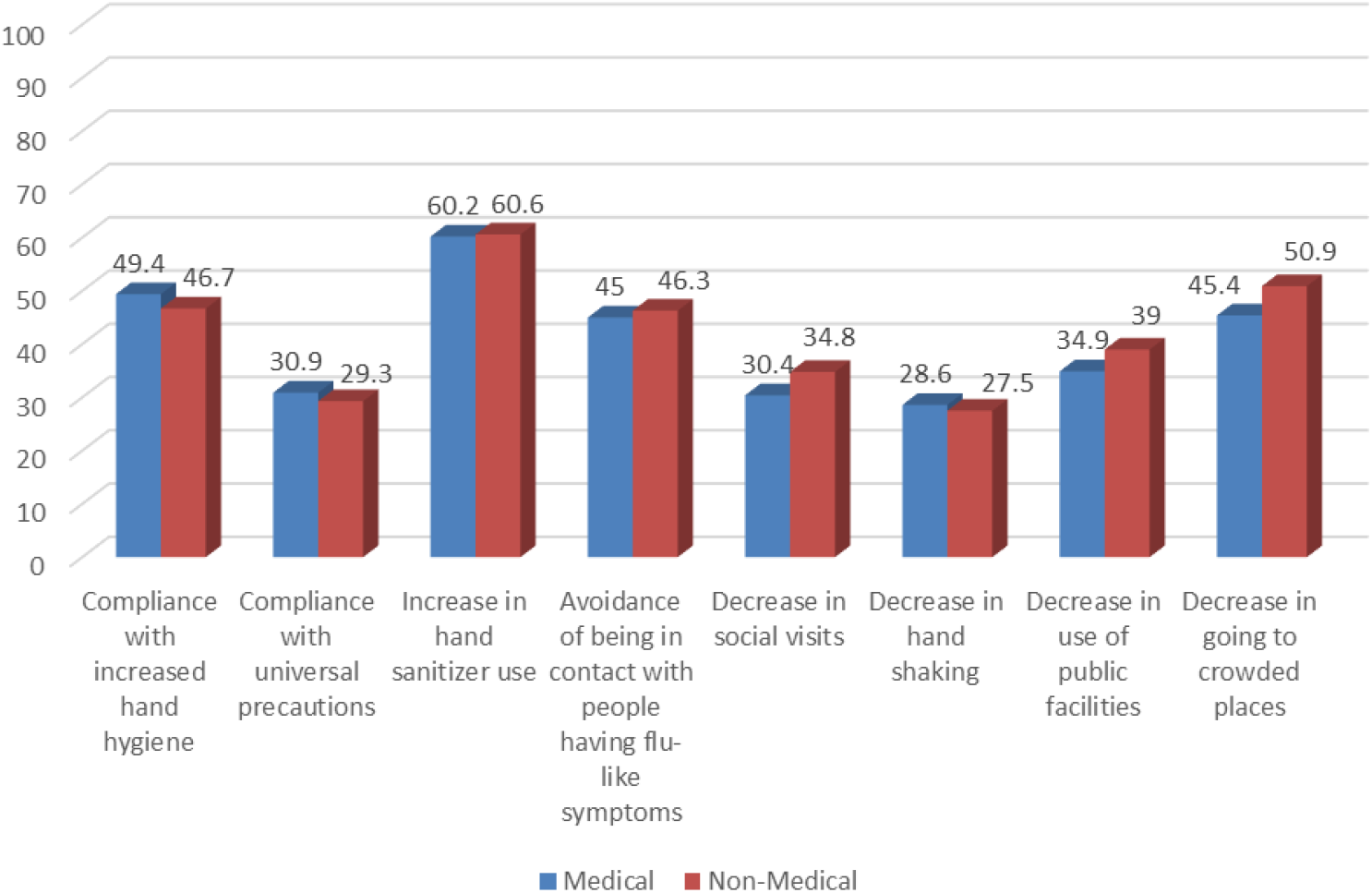
Proportion of Medical & Non-Medical Students Reporting “Very much change” in Hygenic Practices related to COVID-19 outbreaks.

### Knowledge and perception of information on COVID-19

Most students agreed they had heard enough information on the symptoms, prognosis, transmission and prevention of COVID-19. When we compared medical and non-medical students, significantly more medical students reported having heard sufficient information on COVID-19 prognosis, χ^2^ (2, N=1385) = 55.20, p<0.001 and COVID-19 transmission, χ^2^ (2, N=1385) = 17.88, p<0.001. (Table 2). Overall, students had good knowledge of COVID-19 with over 70% of students answering correctly on the COVID-19 knowledge questions with a median score of 5 (from a maximum score of 7). When we compared participant’s’ knowledge scores by field of study, gender and contact with COVID-19, we found that medical students, females and students who had been in contact with COVID-19 cases had significantly higher knowledge scores than their counterparts, as shown in Table 2.

**Table 2:** Perception of sufficient COVID-19 information by field of study and knowledge of COVID-19 by demographic factors.

| I believe I have heard sufficient information about COVID-19, n (%) |  |  |  |  |  |  |
| --- | --- | --- | --- | --- | --- | --- |
|  |  | Strongly disagree/Disagree | Neutral | Agree/ Strongly agree | Chi-Square * | P-value |
| Symptoms | Medical | 94 (11.2) | 87 (10.3) | 660 (78.5) | 2.99 | 0.224 |
|  | Non-medical | 16 (7.8) | 27 (13.2) | 162 (79.0) |  |  |
| Prognosis | Medical | 159 (18.9) | 145 (17.2) | 537 (63.9) | 55.20 | <0.0001 |
|  | Non-medical | 33 (16.1) | 84 (41.0) | 88 (42.9) |  |  |
| Treatment | Medical | 305 (36.3) | 212 (25.2) | 324 (38.5) | 4.74 | 0.094 |
|  | Non-medical | 88 (42.9) | 54 (26.3) | 63 (30.7) |  |  |
| Transmission | Medical | 56 (6.7) | 60 (7.1) | 725 (86.2) | 17.88 | <0.0001 |
|  | Non-medical | 24 (11.7) | 29 (14.1) | 152 (74.1) |  |  |
| Prevention | Medical | 37 (4.4) | 55 (6.5) | 749 (89.1) | 1.88 | 0.391 |
|  | Non-medical | 13 (6.3) | 16 (7.8) | 176 (85.9) |  |  |
| Knowledge score by field of study, gender and contact with COVID-19 patient |  |  |  |  |  |  |
| (Maximum possible 0-7) |  | Median Knowledge Score |  | Mann-Whitney U test |  | P-value |
| Field of Study | Medical | 6 |  | 75060.5 |  | <0.0001 |
|  | Non-Medical | 5 |  |  |  |  |
| Gender | Female | 6 |  | 127281 |  | <0.0001 |
|  | Male | 5 |  |  |  |  |
| Contact with COVID-19 | Yes | 6 |  | 68041 |  | 0.011 |
|  | No | 5 |  |  |  |  |

\*df (Degrees of freedom) = 2

### Student sources of information on COVID-19

Students’ main sources of information were official websites, press releases from the Ministry of Health (MOH) and social media. A statistically significant higher percentage of medical students reported using the WHO website (52%) (*p≤0.001*), MOH (50%) (*p=0.001*), hospital announcements (27%) (*p≤0.001*) and social media (54%) (*p=0.010*) to retrieve their information as displayed in Figure 2.

**Figure 2:**
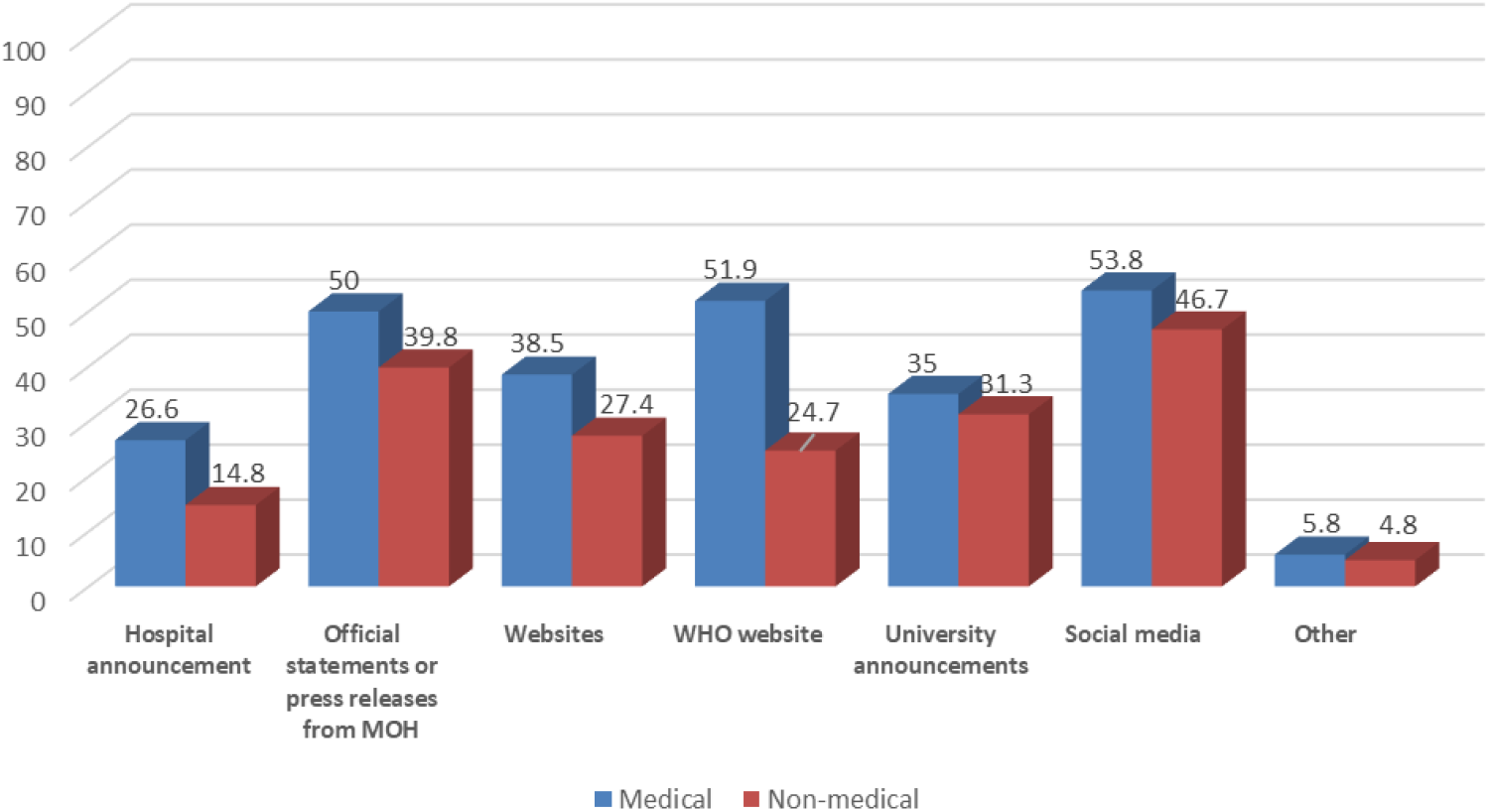
Sources of Information for COVID-19 for Medical and Non-medical Students.

### Levels of anxiety and worry among students

The median GAD-7 scores for medical students during their hospital visits, for all students before introduction of online learning and after online learning were 4, 4 and 3, respectively. When we compared GAD-7 scores for medical students by gender, specialty (dental or medical), contact with COVID-19, rotation type and clinical phase of study, we found that females, dental medicine students, students who had been in contact with COVID-19 patients and students in their clinical phase of study reported statistically significant higher anxiety levels. More students (27%) in low-risk ward rotations reported mild anxiety while the majority of students who reported moderate to severe levels of anxiety (26%) were attending high risk ward rotations χ^2^ (3, N=1385) = 10.92, p=0.027 (Table 3).

**Table 3:** Generalized Anxiety Disorder-7 (GAD-7) Scores (0-21) during hospital visits, before and after online learning by sociodemographic factors.

| GAD-7 score during hospital visits (Medical/dental students) n (%) |  |  |  |  |  |  |  |
| --- | --- | --- | --- | --- | --- | --- | --- |
|  |  | Minimal (0-4) | Mild (5-9) | Moderate/ Severe (10-21) | Chi-Square | df | P-value* |
| Gender | Female | 296 (50.2) | 149 (25.3) | 145 (24.6) | 19.584 | 2 | ≤0.0001 |
|  | Male | 135 (67.5) | 39 (19.5) | 26 (13) |  |  |  |
| Specialty | Medical | 243 (58.1) | 95 (22.7) | 80 (19.2) | 7.535 | 2 | 0.023 |
|  | Dental | 116 (47.5) | 64 (26.2) | 64 (26.2) |  |  |  |
| Contact with COVID-19 | No | 324 (55.9) | 140 (24.1) | 116 (20) | 18.586 | 2 | ≤0.0001 |
|  | Yes | 44 (38.9) | 26 (23) | 43 (38.1) |  |  |  |
| Rotation type | No Rotation | 199 (58.4) | 78 (22.9) | 64 (18.8) | 10.92 | 3 | 0.027 |
|  | High risk** | 18 (66.7) | 2 (7.4) | 7 (25.9) |  |  |  |
|  | Low risk | 142 (48.3) | 79 (26.9) | 73 (24.8) |  |  |  |
| Clinical Phase of study | Pre-clinical | 211 (58.4) | 82 (22.7) | 68 (18.8) | 6.627 | 2 | 0.043 |
|  | Clinical | 148 (49.2) | 77 (25.6) | 76 (25.2) |  |  |  |
| GAD-7 score before online learning (all students) n (%) |  |  |  |  |  |  |  |
| Gender | Female | 376 (47.8) | 216 (27.5) | 194 (24.7) | 22.435 | 2 | ≤0.0001 |
|  | Male | 194 (63.8) | 58 (19.1) | 52 (17.1) |  |  |  |
| Field of Study | Medical | 484 (55.7) | 213 (24.5) | 172 (19.8) | 24.793 | 2 | ≤0.0001 |
|  | Non-medical | 86 (38.9) | 61 (27.6) | 74 (33.5) |  |  |  |
| GAD-7 score after online learning (all students) n (%) |  |  |  |  |  |  |  |
| Gender | Female | 423 (53.8) | 196 (24.9) | 167 (21.2) | 10.788 | 2 | 0.005 |
|  | Male | 197 (64.8) | 58 (19.1) | 49 (16.1) |  |  |  |
| Field of Study | Medical | 546 (62.8) | 194 (22.3) | 129 (14.8) | 81.90 | 2 | ≤0.0001 |
|  | Non-medical | 74 (33.5) | 60 (27.1) | 87 (39.4) |  |  |  |
| Change in GAD-7 scores before and after online learning n (%) |  |  |  |  |  |  |  |
|  |  | Before>After | Before=After | Before <After |  |  |  |
| Gender | Female | 184 (23.4) | 479 (60.9) | 123 (15.6) | 12.224 | 2 | 0.0002 |
|  | Male | 47 (15.5) | 219 (72) | 38 (12.5) |  |  |  |
| Specialty | Medical | 109 (18.2) | 405 (67.5) | 86 (14.3) | 36.383 | 4 | ≤0.0001 |
|  | Dental | 77 (28.6) | 171 (63.6) | 21 (7.8) |  |  |  |
|  | Non-medical | 45 (20.4) | 122 (55.2) | 54 (24.4) |  |  |  |
| Field of Study | Medical | 186 (21.4) | 576 (66.3) | 107 (12.3) | 20.994 | 2 | ≤0.0001 |
|  | Non-medical | 45 (20.4) | 122 (55.2) | 54 (24.4) |  |  |  |
| Contact with COVID-19 | No | 200 (20.9) | 612 (64) | 144 (15.1) | 0.722 | 2 | 0.697 |
|  | Yes | 31 (23.1) | 86 (64.2) | 17 (12.7) |  |  |  |

\*Significant at $p < 0.05$ \*\*High risk rotation includes ICU, ER, OR, Isolation wards

Overall, females reported higher levels of anxiety in GAD-7, both before and after online learning. The medical students reported higher levels of anxiety before online learning in comparison to non-medical students (Table 3).

Interestingly, anxiety levels significantly decreased for females and for medical students after switching to the online learning, with χ^2^ (2, N=1385) = 12.22, p<0.001 and χ^2^ (2, N=1385) = 20.99, p<0.001, respectively, with a higher percentage of decreased anxiety among dental students; whilst non-medical students reported higher levels of anxiety after the introduction of online learning (χ^2^ (4, N=1385) = 36.38, p<0.001) (Table 3).

The majority of students (73%) reported being worried about transmitting COVID-19 to one of their family members or friends, while 65% of them were worried about catching the virus themselves. Almost half (49%) of medical students reported being worried about transmitting COVID-19 to others and were less likely (29%) to worry about catching it themselves, in comparison to non-medical students (35%), although the differences were not statistically significant (χ^2^ (2, N=1385) = 2.85, p=0.24). Most students (61%) agreed that the public fear was justifiable and (46%) of students did not perceive it as dysfunctional.

A cumulative odds ordinal logistic regression with proportional odds was performed to determine predictors of anxiety levels during the different time points. The effect of gender, specialty, contact with COVID-19, rotation type, clinical phase of study and knowledge score on the GAD-7 score during hospital visits was modelled. We included knowledge score in the regression model because we found a positive, but weak, correlation between knowledge score and anxiety score using Spearman’s correlation (r_s_=0.086, p=0.015). The odds of females having higher anxiety scores during hospital visits was 2.02 (95% CI, 1.41 to 2.91) times that for males, and medical students reported lower anxiety levels in comparison to dental medicine students (OR=0.61, 95% CI, 0.45 to 0.84). Furthermore, being in a rotation and demonstrating higher knowledge scores predicted higher GAD-7 scores during hospital visits (Table 4). Similarly, gender, field of study and knowledge score predicted higher GAD-7 scores before online learning, but only gender and field of study predicted GAD-7 scores after switching to online learning (Table 4).

**Table 4:** Ordinal logistic regression model for factors predicting anxiety GAD-7 score.

|  | **n (%)** | **b** | **SE(b)** | **P-Value** | **OR [95% CI]** |
| --- | --- | --- | --- | --- | --- |
| *GAD-7 score during hospital visits* |  |  |  |  |  |
| Gender Female | 492 (74.3) | 0.71 | 0.19 | ≤0.001 | 2.02 [1.41-2.91] |
| Male<sup>a</sup> | 170 (25.7) | - | - | - | 1 |
| Specialty Medical | 418 (63.1) | -0.49 | 0.16 | 0.002 | 0.61 [0.45-0.84] |
| Dental<sup>a</sup> | 244 (36.9) | - | - | - | 1 |
| Contact with COVID-19 |  |  |  |  |  |
| No | 552 (83.4) | -0.77 | 0.21 | ≤0.001 | 0.46 [0.31- 0.69] |
| Yes<sup>a</sup> | 110 (16.6) | - | - | - | 1 |
| In Rotation |  |  |  |  |  |
| No | 341 (51.5) | -0.09 | 0.22 | 0.672 | 0.91 [0.60-1.40] |
| Yes<sup>a</sup> | 321 (48.5) | - | - | - | 1 |
| Clinical phase of study |  |  |  |  |  |
| Pre-clinical | 361 (54.5) | -0.04 | 0.22 | 0.856 | 0.96 [0.62-1.47] |
| Clinical<sup>a</sup> | 301(45.5) | - | - | - | 1 |
| Knowledge score | 662 | 0.18 | 0.08 | 0.024 | 1.19 [1.02-1.40] |
| **-2Log-Likelihood 424.53 Likelihood Ratio Chi-Square 48.790 (df=6, p-value≤0.001)** |  |  |  |  |  |
| *GAD-7 score before online learning* |  |  |  |  |  |
| Gender Female | 786 (72.1) | 0.61 | 0.14 | ≤0.001 | 1.85 [1.41-2.41] |
| Male<sup>a</sup> | 304 (27.9) | - | - | - | 1 |
| Field of study |  |  |  |  |  |
| Medical | 869 (79.7) | -0.87 | 0.15 | ≤0.001 | 0.42 [0.31-0.56] |
| Non-medical<sup>a</sup> | 221 (20.3) | - | - | - | 1 |
| Knowledge score | 1090 | 0.18 | 0.06 | 0.002 | 1.20 [1.07-1.34] |
| **-2Log-Likelihood 208.599 Likelihood Ratio Chi-Square 58.130 (df=3, p-value≤0.001)** |  |  |  |  |  |
| *GAD score after online learning* |  |  |  |  |  |
| Gender Female | 786 (72.1) | 0.541 | 0.141 | ≤0.001 | 1.72 [1.30-2.26] |
| Male<sup>a</sup> | 304 (27.9) | - | - | - | 1 |
| Field of study |  |  |  |  |  |
| Medical | 869 (79.7) | -1.389 | 0.152 | ≤0.001 | 0.25 [0.189-0.34] |
| Non-medical<sup>a</sup> | 221 (20.3) | - | - | - | 1 |
| Knowledge score | 1090 | 0.090 | 0.058 | 0.122 | 1.10 [0.98-1.23] |
| **-2Log-Likelihood 185.344 Likelihood Ratio Chi-Square 95.435 (df=3, p-value≤0.001)** |  |  |  |  |  |

<sup>a</sup> reference group, *b* parameter estimate, *SE* Std Error, *OR* Odds Ratio, *CI* Confidence Interval

## Discussion

This study has revealed that the COVID-19 pandemic has impacted on anxiety levels among university students in the UAE, with almost half of students reporting mild to moderate/severe anxiety levels. The effect of COVID-19 on the global community overall has been considerably significant, causing fear, anxiety and worry, particularly due to uncertainty of the prognosis of the disease, changes in societies lifestyles, lockdown restrictions and educational disruptions. The impact of COVID-19 on university students is particularly burdensome due to the perceived effect of the virus on their studies; and with medical students specifically it could be due to the interdisciplinary nature of their training and the potential proximity of being exposed to the virus during their clinical studies [17]

Overall, students in our study demonstrated good knowledge of COVID-19 and reported using reliable sources such as official statements and press releases from the ministry of health and the WHO website. Although the use of these sources was higher among medical students, social media was the main source of information for both medical and non-medical students. This finding is consistent with the literature and emphasizes the role that social media can play in risk perception and dissemination of reliable information during a pandemic such as COVID-19 [18-21]. However, other studies have also reported that young people tend to obtain a large amount of information from social media which can easily be a trigger for stress and anxiety [22].

Good knowledge may also explain students’ compliance with hygienic practices in our study. Most students reported significant change in their hygienic behavior since the COVID-19 outbreak, particularly for increased hand hygiene, avoiding crowded places and avoiding being in contact with people with flu like symptoms. However, even though levels of knowledge were significantly higher among medical students, compliance with hygienic practices were similar for both groups. Furthermore, we found significant positive correlations between changes in hygienic behavior and increased levels of anxiety. More than half of students who reported changed hygienic practices reported higher levels of anxiety, which is consistent with recent literature indicating that people who were more anxious about COVID-19 were also more engaged with regular hand hygiene and social distancing behaviors [23].

There is increasing evidence that the number of growing cases of COVID-19 globally and within the UAE is causing public worry and concern[24, 25] and in the absence of vaccines and effective treatment, government authorities have introduced rules and restrictions[26]. Compliance and adherence with these restrictions has been found to differ amongst different age groups and populations, with less acceptance of these restrictions being reported amongst younger age-groups [27]. However, in our study, the majority of students did not consider the strict measures undertaken by the healthcare authorities as dysfunctional or not required. The students were worried about transmitting COVID-19 to their family members more than they were worried about catching the virus themselves, therefore, indicating a beneficial sense of social responsibility during such infectious disease outbreak. Similar concerns were reported amongst healthcare workers in Hong Kong and Canada during the SARS outbreak [28, 29].

The results from the current study confirm that anxiety levels due to COVID-19 among university students are high, ranging from mild to severe, especially amongst females, which is in accordance with previous research [22, 30, 31]. The majority of students in our study were females, which reflects the gender imbalance in higher education within the UAE. More than two thirds of students reported mild anxiety levels and one third of students reported severe anxiety. What makes this study unique, is that medical and non-medical students were compared, and within the medical student group, we compared medical and dental students. Furthermore, this study assessed anxiety levels at three different time points: during hospital visits for medical and dental students, before online learning and after switching to the online learning for all students. Among the medical/dental group, students in their clinical phase of study and who had rotations in high risk wards or who had been in contact with COVID-19 patients reported significantly higher (moderate to severe) levels of anxiety, demonstrating that high risk perception of COVID-19 may contribute to higher levels of anxiety. Medical/dental students continued attending the hospitals at the early onset of COVID-19 when fear and worry associated with the outbreak would have been at their peak and before lockdown restrictions were in place, which could also explain their high compliance with infection control measures. Medical students high risk perceptions associated with attending hospitals during infectious disease outbreaks has been reportedly associated with higher levels of anxiety [12, 21]. However, when we compared between medical and dental students, dental students reported higher levels of anxiety and these remained significant with further multivariate analysis. Previous studies have established that medical and dental programs are highly competitive and students generally experience high levels of stress during their training [32, 33]. Yet, dental students suffer from greater levels of perceived stress than medical students due to their role as providers of care and earlier exposure to patients in the dental clinics [34]. Furthermore, dental students are in very close proximity to patients and are dealing directly with patients’ dental care during a potentially highly transmissible respiratory virus. This may explain the higher levels of anxiety among dental students in our study and the overall decrease in anxiety among all medical students following the introduction of online learning. Psychological support should be tailored to each students’ needs and incorporated into the online remote curriculum. Screening university students on a regular basis with tools such as the GAD-7 could help faculty in the early identification of highly anxious students and guide students to receive help from targeted interventions that promote psychological well-being, offer counselling, mental health support and coping mechanisms[35].

Non-medical students reported higher levels of anxiety before and after online learning in comparison to medical students and whilst medical student anxiety levels decreased following the introduction of online learning, non-medical student’s anxiety levels increased. This may be due to several factors, including medical students possibly being more familiar with the use of online learning platforms, being distant from the perceived risk of COVID-19, or due to the variable sources of information about the pandemic among both groups. Knowledge of the virus might reduce students fears and anxiety while inadequate understanding of COVID-19, its prognosis, transmission and control measures might contribute to negative implications and fear of the unknown [29] hence explaining the higher anxiety levels in non-medical students in our study.

### Limitations

Despite the findings of this study, we acknowledge that it has several limitations. Firstly, the use of convenience sampling and its descriptive nature through an online survey may not allow the generalization of results. However, considering the need for a rapid method to assess stress and anxiety in a vulnerable population during a rapidly evolving infectious disease outbreak, the use of an online survey serves as a promising method for quick results [36]. Additionally, responses were collected from four different emirates across the UAE with good response rate allowing for a certain element of representation. Secondly, the nature of self-reported data in the survey may lead to response biases specifically for hygienic practices where students may provide socially desirable responses and self-reported levels of anxiety, stress and worry which may not always be as accurate as being assessed by a mental health professional. However, despite these limitations, this study provides important baseline information which will inform further research and public health interventions in this area.

### Conclusion

To the best of our knowledge, this is the first study to provide important information on the initial response and anxiety levels in university students across the UAE immediately following the period COVID-19 was declared a global pandemic. More than half of university students reported mild to severe anxiety levels with a quarter of students reporting severe anxiety. Specifically, medical students reported higher levels of anxiety during their clinical rotations which decreased with the introduction of online learning, yet, non-medical student’s anxiety levels increased with online learning. The findings from our study can be used at the government and university level to develop effective screening strategies and to formulate interventions that improve mental health of students. Such strategies will reduce unnecessary stress and anxiety among university students, as well as build psychological resilience during the COVID-19 pandemic or any other public health emergencies in the future.

## Data Availability

The raw data supporting the conclusions of this article will be made available by the authors, without undue reservation.

## Author Contributions

RH, BS, FS, WK and MT conceived, designed and initiated the study. RK, EH, NA, FM contributed to the planning and implementation of the study. AH analyzed survey data. BS, AH, RH interpreted the results. BS drafted the manuscript with input from RH, AH, FS, MT and QH. All authors read and approved the final version of the manuscript.

## Acknowledgements

The authors would like to thank all students who participated in the study.

## Supplementary Material

The survey used to collect data for this study is provided as supplementary material.

## References

1. Paules CI, Marston HD, Fauci AS. Coronavirus Infections-More Than Just the Common Cold. Jama. 2020. Epub 2020/01/24. doi: 10.1001/jama.2020.0757. PubMed PMID: 31971553.

2. WHO. World Health Organisation Director General’s opening remarks at the media briefing on COVID-19, 11th March 2020. https://www.whoint/dg/speeches/detail/who-director-general-s-opening-remarks-at-the-media-briefing-on-covid-19---11-march-2020. 30–3-2020.

3. WHO. World Health Organization. Coronavirus disease (COVID-19) outbreak. https://covid19whoint/. 3–5-2020.

4. DOH. Department of Health (UAE).. https://www.dohgovae/covid-19. 3–5-2020.

5. Al Saadi T, Zaher Addeen S, Turk T, Abbas F, Alkhatib M. Psychological distress among medical students in conflicts: a cross-sectional study from Syria. BMC medical education. 2017;17(1):173. Epub 2017/09/22. doi: 10.1186/s12909-017-1012-2. PubMed PMID: 28931387; PubMed Central PMCID: PMCPMC5607487.

6. Quek TT, Tam WW, Tran BX, Zhang M, Zhang Z, Ho CS, et al. The Global Prevalence of Anxiety Among Medical Students: A Meta-Analysis. International journal of environmental research and public health. 2019;16(15). Epub 2019/08/03. doi: 10.3390/ijerph16152735. PubMed PMID: 31370266; PubMed Central PMCID: PMCPMC6696211.

7. Trivate T, Dennis AA, Sholl S, Wilkinson T. Learning and coping through reflection: exploring patient death experiences of medical students. BMC medical education. 2019;19(1):451. Epub 2019/12/06. doi: 10.1186/s12909-019-1871-9. PubMed PMID: 31801494; PubMed Central PMCID: PMCPMC6894273.

8. Kim S, Kim S. Exploring the Determinants of Perceived Risk of Middle East Respiratory Syndrome (MERS) in Korea. International journal of environmental research and public health. 2018;15(6). Epub 2018/06/06. doi: 10.3390/ijerph15061168. PubMed PMID: 29867054; PubMed Central PMCID: PMCPMC6025578.

9. Al Ghobain M, Aldrees T, Alenezi A, Alqaryan S, Aldabeeb D, Alotaibi N, et al. Perception and Attitude of Emergency Room Resident Physicians toward Middle East Respiratory Syndrome Outbreak. Emergency medicine international. 2017;2017:6978256. Epub 2017/05/11. doi: 10.1155/2017/6978256. PubMed PMID: 28487774; PubMed Central PMCID: PMCPMC5402244.

10. Goulia P, Mantas C, Dimitroula D, Mantis D, Hyphantis T. General hospital staff worries, perceived sufficiency of information and associated psychological distress during the A/H1N1 influenza pandemic. BMC infectious diseases. 2010;10:322. Epub 2010/11/11. doi: 10.1186/1471-2334-10-322. PubMed PMID: 21062471; PubMed Central PMCID: PMCPMC2990753.

11. Al-Rabiaah A, Temsah MH, Al-Eyadhy AA, Hasan GM, Al-Zamil F, Al-Subaie S, et al. Middle East Respiratory Syndrome-Corona Virus (MERS-CoV) associated stress among medical students at a university teaching hospital in Saudi Arabia. Journal of infection and public health. 2020. Epub 2020/02/01. doi: 10.1016/j.jiph.2020.01.005. PubMed PMID: 32001194; PubMed Central PMCID: PMCPMC7102651.

12. Wong TW, Gao Y, Tam WWS. Anxiety among university students during the SARS epidemic in Hong Kong. Stress and health. 2007;23(1):31–5. doi: 10.1002/smi.1116.

13. SurveyMonkey. San Mateo. CA.

14. Spitzer RL, Kroenke K, Williams JB, Lowe B. A brief measure for assessing generalized anxiety disorder: the GAD-7. Archives of internal medicine. 2006;166(10):1092–7. Epub 2006/05/24. doi: 10.1001/archinte.166.10.1092. PubMed PMID: 16717171.

15. Kroenke K, Spitzer RL, Williams JB, Monahan PO, Lowe B. Anxiety disorders in primary care: prevalence, impairment, comorbidity, and detection. Annals of internal medicine. 2007;146(5):317–25. Epub 2007/03/07. doi: 10.7326/0003-4819-146-5-200703060-00004. PubMed PMID: 17339617.

16. IBM Corp. Released 2017. IBM SPSS Statistics for Windows VA, NY: IBM Corp.

17. Rastegar Kazerooni A, Amini M, Tabari P, Moosavi M. Peer mentoring for medical students during COVID-19 pandemic via a social media platform. Medical education. 2020. Epub 2020/05/01. doi: 10.1111/medu.14206. PubMed PMID: 32353893.

18. Albarrak AI, Mohammed R, Al Elayan A, Al Fawaz F, Al Masry M, Al Shammari M, et al. Middle East Respiratory Syndrome (MERS): Comparing the knowledge, attitude and practices of different health care workers. Journal of infection and public health. 2019. Epub 2019/08/23. doi: 10.1016/j.jiph.2019.06.029. PubMed PMID: 31431424; PubMed Central PMCID: PMCPMC7102554.

19. Karasneh R, Al-Azzam S, Muflih S, Soudah O, Hawamdeh S, Khader Y. Media’s effect on shaping knowledge, awareness risk perceptions and communication practices of pandemic COVID-19 among pharmacists. Research in social & administrative pharmacy: RSAP. 2020. Epub 2020/04/29. doi: 10.1016/j.sapharm.2020.04.027. PubMed PMID: 32340892; PubMed Central PMCID: PMCPMC7179508.

20. Khan MU, Shah S, Ahmad A, Fatokun O. Knowledge and attitude of healthcare workers about Middle East Respiratory Syndrome in multispecialty hospitals of Qassim, Saudi Arabia. BMC public health. 2014;14:1281. Epub 2014/12/17. doi: 10.1186/1471-2458-14-1281. PubMed PMID: 25510239; PubMed Central PMCID: PMCPMC4300996.

21. Yang S, Cho SI. Middle East respiratory syndrome risk perception among students at a university in South Korea, 2015. American journal of infection control. 2017;45(6):e53-e60. Epub 2017/04/08. doi: 10.1016/j.ajic.2017.02.013. PubMed PMID: 28385465; PubMed Central PMCID: PMCPMC7115287.

22. Qiu J, Shen B, Zhao M, Wang Z, Xie B, Xu Y. A nationwide survey of psychological distress among Chinese people in the COVID-19 epidemic: implications and policy recommendations. General psychiatry. 2020;33(2):e100213. Epub 2020/03/28. doi: 10.1136/gpsych-2020-100213. PubMed PMID: 32215365; PubMed Central PMCID: PMCPMC7061893.

23. Harper CA, Satchell LP, Fido D, Latzman RD. Functional Fear Predicts Public Health Compliance in the COVID-19 Pandemic. International journal of mental health and addiction. 2020:1–14. Epub 2020/04/30. doi: 10.1007/s11469-020-00281-5. PubMed PMID: 32346359; PubMed Central PMCID: PMCPMC7185265.

24. Bao Y, Sun Y, Meng S, Shi J, Lu L. 2019-nCoV epidemic: address mental health care to empower society. Lancet (London, England). 2020;395(10224):e37-e8. Epub 2020/02/12. doi: 10.1016/s0140-6736(20)30309-3. PubMed PMID: 32043982; PubMed Central PMCID: PMCPMC7133594.

25. Rajkumar RP. COVID-19 and mental health: A review of the existing literature. Asian journal of psychiatry. 2020;52:102066. Epub 2020/04/18. doi: 10.1016/j.ajp.2020.102066. PubMed PMID: 32302935; PubMed Central PMCID: PMCPMC7151415.

26. Ryan BJ, Coppola D, Canyon DV, Brickhouse M, Swienton R. COVID-19 Community Stabilization and Sustainability Framework: An Integration of the Maslow Hierarchy of Needs and Social Determinants of Health. Disaster medicine and public health preparedness. 2020:1–16. Epub 2020/04/22. doi: 10.1017/dmp.2020.109. PubMed PMID: 32314954.

27. Zettler I, Christoph Schild, Lau Lilleholt, and Robert Böhm. Individual Differences in Accepting Personal Restrictions to Fight the COVID-19 Pandemic: Results from a Danish Adult Sample. PsyArXiv. 2020;Preprint. Epub 23 Mar 2020. doi: doi:10.31234/osf.io/pkm2a.

28. Maunder RG, Lancee WJ, Balderson KE, Bennett JP, Borgundvaag B, Evans S, et al. Long-term psychological and occupational effects of providing hospital healthcare during SARS outbreak. Emerging infectious diseases. 2006;12(12):1924–32. Epub 2007/03/01. doi: 10.3201/eid1212.060584. PubMed PMID: 17326946; PubMed Central PMCID: PMCPMC3291360.

29. Wong JG, Cheung EP, Cheung V, Cheung C, Chan MT, Chua SE, et al. Psychological responses to the SARS outbreak in healthcare students in Hong Kong. Medical teacher. 2004;26(7):657–9. Epub 2005/03/15. doi: 10.1080/01421590400006572. PubMed PMID: 15763860.

30. Liu N, Zhang F, Wei C, Jia Y, Shang Z, Sun L, et al. Prevalence and predictors of PTSS during COVID-19 outbreak in China hardest-hit areas: Gender differences matter. Psychiatry research. 2020;287:112921. Epub 2020/04/03. doi: 10.1016/j.psychres.2020.112921. PubMed PMID: 32240896; PubMed Central PMCID: PMCPMC7102622 of interest.

31. Wang C, Pan R, Wan X, Tan Y, Xu L, Ho CS, et al. Immediate Psychological Responses and Associated Factors during the Initial Stage of the 2019 Coronavirus Disease (COVID-19) Epidemic among the General Population in China. International journal of environmental research and public health. 2020;17(5). Epub 2020/03/12. doi: 10.3390/ijerph17051729. PubMed PMID: 32155789; PubMed Central PMCID: PMCPMC7084952.

32. Aboalshamat K, Hou XY, Strodl E. Psychological well-being status among medical and dental students in Makkah, Saudi Arabia: a cross-sectional study. Medical teacher. 2015;37 Suppl 1:S75–81. Epub 2015/02/05. doi: 10.3109/0142159x.2015.1006612. PubMed PMID: 25649101.

33. Ahmad FA, Karimi AA, Alboloushi NA, Al-Omari QD, AlSairafi FJ, Qudeimat MA. Stress Level of Dental and Medical Students: Comparison of Effects of a Subject-Based-Curriculum versus a Case-Based Integrated Curriculum. Journal of dental education. 2017;81(5):534–44. Epub 2017705704. doi: 10.218157jde.016.026. PubMed PMID: 28461630.

34. Murphy RJ, Gray SA, Sterling G, Reeves K, DuCette J. A comparative study of professional student stress. Journal of dental education. 2009;73(3):328–37. Epub 2009/03/18. PubMed PMID: 19289722.

35. Roberto A, Almeida A. [Mental health of students of medicine: exploraty study in the Universidade da Beira Interior]. Acta medica portuguesa. 2011;24 Suppl 2:279–86. Epub 2012/08/08. PubMed PMID: 22849913.

36. Geldsetzer P. Use of Rapid Online Surveys to Assess People’s Perceptions During Infectious Disease Outbreaks: A Cross-sectional Survey on COVID-19. Journal of medical Internet research. 2020;22(4):e18790. Epub 2020/04/03. doi: 10.2196718790. PubMed PMID: 32240094; PubMed Central PMCID: PMCPMC7124956.

